# A high protein meal does not change autophagy in human blood

**DOI:** 10.1101/2024.10.28.24316074

**Authors:** S Singh, C Fourrier, K Hattersley, LK Hein, J Gore, LK Heilbronn, J Bensalem, TJ Sargeant

**Affiliations:** Lysosomal Health in Ageing, Lifelong Health Theme, South Australian Health and Medical Research Institute (SAHMRI), Adelaide, Australia; Faculty of Health and Medical Sciences, The University of Adelaide, Adelaide,, Australia; SAHMRI Clinical Trials Platform, Adelaide, Australia; Adelaide Medical School, The University of Adelaide, Adelaide, Australia; Lifelong Health Theme, South Australian Health and Medical Research Institute (SAHMRI), Adelaide, Australia

## Abstract

Autophagy is a catabolic quality control pathway that has been linked to neurodegenerative disease, atherosclerosis and ageing, and can be modified by nutrient availability in preclinical models. Consequently, there is immense public interest in stimulating autophagy in people. However, progress has been hampered by the lack of techniques to measure human autophagy. As a result, several key concepts in the field, including nutritional modulation of autophagy, have yet to be validated in humans. We conducted a single arm pre-post study in 42 healthy individuals, to assess whether an acute nutritional intervention could modify autophagy in humans. Two blood samples were collected per participant: after a 12 h overnight fast and 1 h post-consumption of a high protein meal. Autophagy turnover was assessed using a physiologically relevant measure of autophagic flux in peripheral blood mononuclear cells. A lysosomal inhibitor was added directly to whole blood, with the resulting build-up of autophagy marker LC3B-II designated as flux, and measured quantitatively via ELISA. Notably, consumption of a high protein meal had no impact on autophagy, with no differences between overnight fasting and postprandial autophagic flux. We observed sexual dimorphism in autophagy, with females having higher autophagic flux compared to males (p = 0.0031). Exploratory analyses revealed sex-specific correlations between autophagy, insulin and glucose signalling. Importantly, our findings show that an acute nutritional intervention (overnight fasting followed by consumption of a protein-rich meal) does not change autophagic flux in humans, highlighting the need to conduct further autophagy studies in humans.

## Research Letter

Autophagy is a catabolic quality control pathway activated by a myriad of stressors – such as starvation or the presence of pathogenic protein aggregates – to mediate the lysosomal turnover of material and restore cellular homeostasis. Defects in autophagy have been linked to neurodegeneration, atherosclerosis, and ageing [1]. Activating autophagy extends both life- and health span in preclinical models [1]. Consequently, there is immense interest in stimulating autophagy in people. However, progress has been hampered by the lack of techniques to measure human autophagy. The gold-standard is to use a drug to block lysosomal degradation and quantify the resulting build-up of a key autophagy protein LC3B-II as ‘flux’, i.e. the amount of material that would have been turned over in the ensuing time. While this is possible using cultured human peripheral blood mononuclear cells (PBMCs) *ex vivo* [2], it alters their extracellular nutrient composition – a profound autophagy regulator – and does not accurately reflect physiological autophagy. Recently, we developed a test to measure autophagic flux in whole blood [3], but so far it has only been used in observational studies.

Nutrient restriction is a universal activator of autophagy; conversely, high protein intake dampens autophagic flux in preclinical models. We conducted a single arm pre-post study in 42 healthy individuals to assess whether consumption of a high protein meal could acutely modify autophagy in humans (Supplemental Methods). We hypothesised that autophagic flux would be high in overnight fasted individuals, and supressed after consumption of a protein-rich meal. Two blood samples were collected per participant: after a 12 h overnight fast and 1 h post-consumption of a high protein meal (Figure 1A). Autophagic turnover was assessed by adding a lysosomal inhibitor to whole blood, with the resulting build-up of the autophagy marker LC3B-II designated as ‘flux’ and measured quantitatively via ELISA. The primary outcome of this study was to assess the change (ΔLC3B-II flux) between fasted and post-prandial autophagic flux.

**Figure 1.**
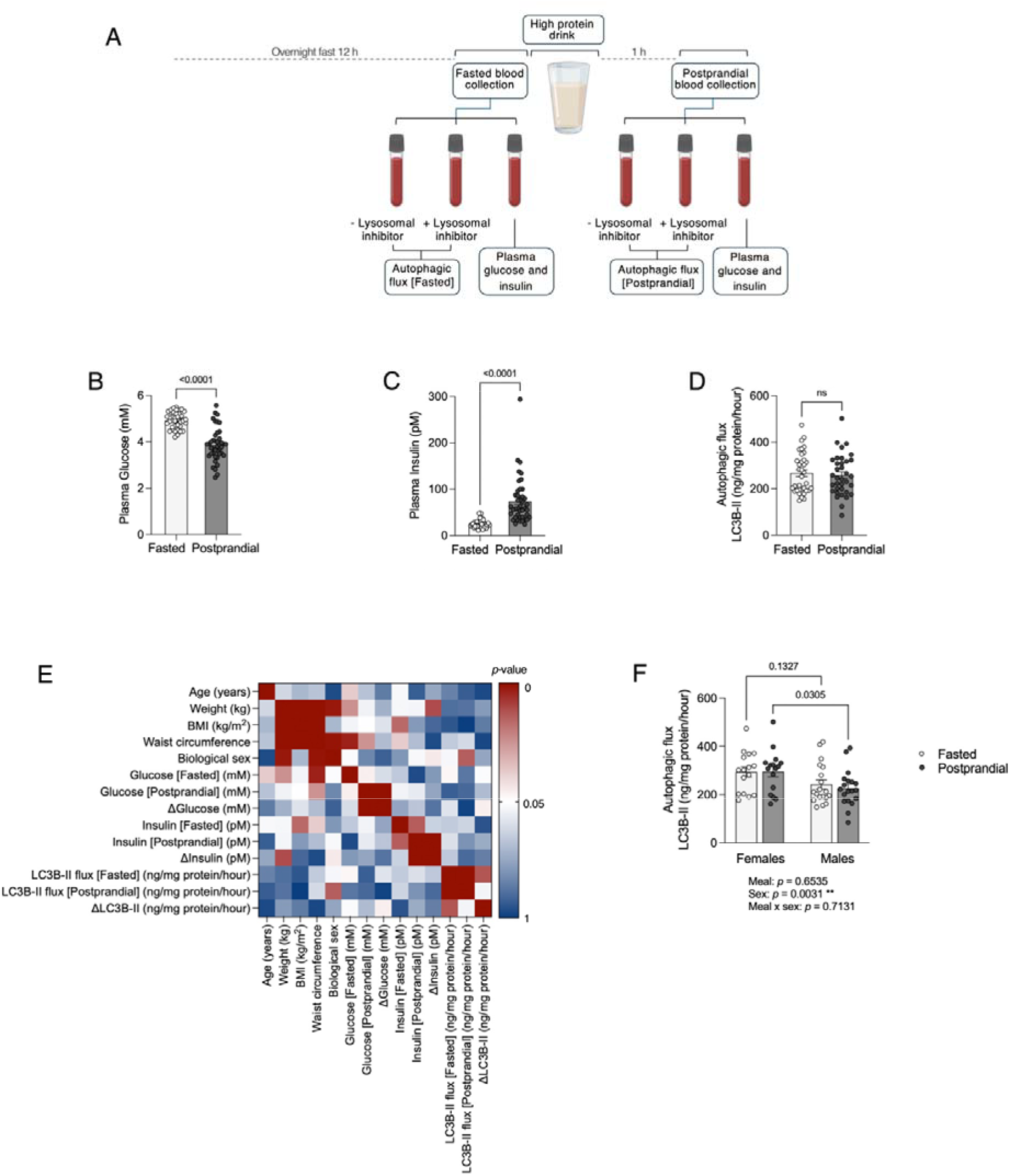
A high protein meal does not alter autophagic flux in the blood of fasted, healthy individuals. **(A)** Schematic of study design. Fasted and post-prandial plasma **(B)** glucose (paired t-test, n=41-42), **(C)** insulin (Wilcoxon test, n=41-42) and **(D)** autophagic flux (Wilcoxon test, n=35). **(E)** Heatmap showing p-values of Spearman’s correlations between blood measurements and baseline characteristics. **(F)** Two-way ANOVA of autophagic flux with sex and meal status as factors (n=17-18).

Participant characteristics are presented in Supplemental Table 1. Plasma glucose decreased and insulin increased 1 h after consumption of a whey protein drink (Figure 1B,C), indicating we were within the window to detect post-prandial effects in the blood. Strikingly, consumption of a protein-rich drink had no impact on autophagy: there were no differences between fasting and post-prandial autophagic flux (Figure 1D). Spearman’s analyses revealed a correlation between autophagic flux and biological sex (Figure 1E). We therefore stratified autophagy by sex and found that females had greater levels of flux compared to males (Figure 1F, p = 0.0031).

Here, we demonstrate that in contrast to animal models which show that autophagy is down-regulated upon food intake [2,4], autophagic flux in human blood remains unchanged 1 h post-consumption of a protein-rich meal. Using nutrition to modulate autophagy is of considerable interest to both the general public and scientific community – there are already products on the market claiming to activate autophagy in people via this pathway. Previous work has shown that LC3B-II decreased in human muscle 90 min after consuming a mixed macronutrient meal containing 22 g protein [5], but the study solely examined steady-state levels of LC3B-II, which does not provide information on autophagic turnover. Others have reported increased autophagic flux in PBMCs isolated after 24 h starvation in healthy human volunteers [2], however measurement was performed *ex vivo* using nutrient-rich culture medium. This is suboptimal, since the heightened autophagic flux in starved cells can disappear in as little as 10 min following restimulation with nutrients. To our knowledge, this study is the first to examine autophagic flux in physiologically intact human tissue in response to a nutritional intervention. We show that, contrary to the widespread notion that fasting activates autophagy, there is no difference between overnight fasting and post-prandial autophagic flux in humans. However, we did observe sexual dimorphism in autophagic flux, with females having higher levels compared to males.

Our work comes with some caveats. LC3B lipidation, the readout of our assay, can also occur during non-canonical autophagy, and its contribution to our autophagy measurements is currently unknown. While we observed expected changes in plasma glucose and insulin in participants 1 h post-consumption of a high protein drink, this sampling time-point may not have been optimal for detecting changes to autophagic flux. Further, tissue-specific autophagy responses have been well-documented in the literature [1]. It is possible that our nutritional intervention changed autophagic flux in other tissues such as liver or muscle, but not in PBMCs. This is an inherent limitation of human autophagy measurement because, at present, blood is the most accessible source of human tissue amenable to flux assays.

We have previously observed an age-associated increase in PBMC flux in an older, pre-diabetic cohort [6], as well as reduction in flux upon the addition of exogenous insulin and leucine to blood [3]. This suggests that, at least in some scenarios, our methodology is equipped to detect changes in autophagy.

In summary, our results show that autophagic flux in humans remains unchanged after consumption of a protein-rich meal. Importantly, this challenges the assumption that an acute nutritional stimulus can alter autophagy in people. It remains to be seen whether longer-term interventions such as intermittent fasting or caloric restriction change flux in humans. Given that autophagy has been linked to several pathologies [1], augmenting this pathway is of immense clinical interest. Our findings highlight discrepancies between preclinical and clinical modulation of autophagy, underscoring the need for human autophagy research to answer the question – how can we boost autophagy in people?

## Data Availability

All data produced in the present study are available upon reasonable request to the authors

