## Supplemental for "A high protein meal does not change autophagy in human blood"

**SUPPLEMENTARY INFORMATION**

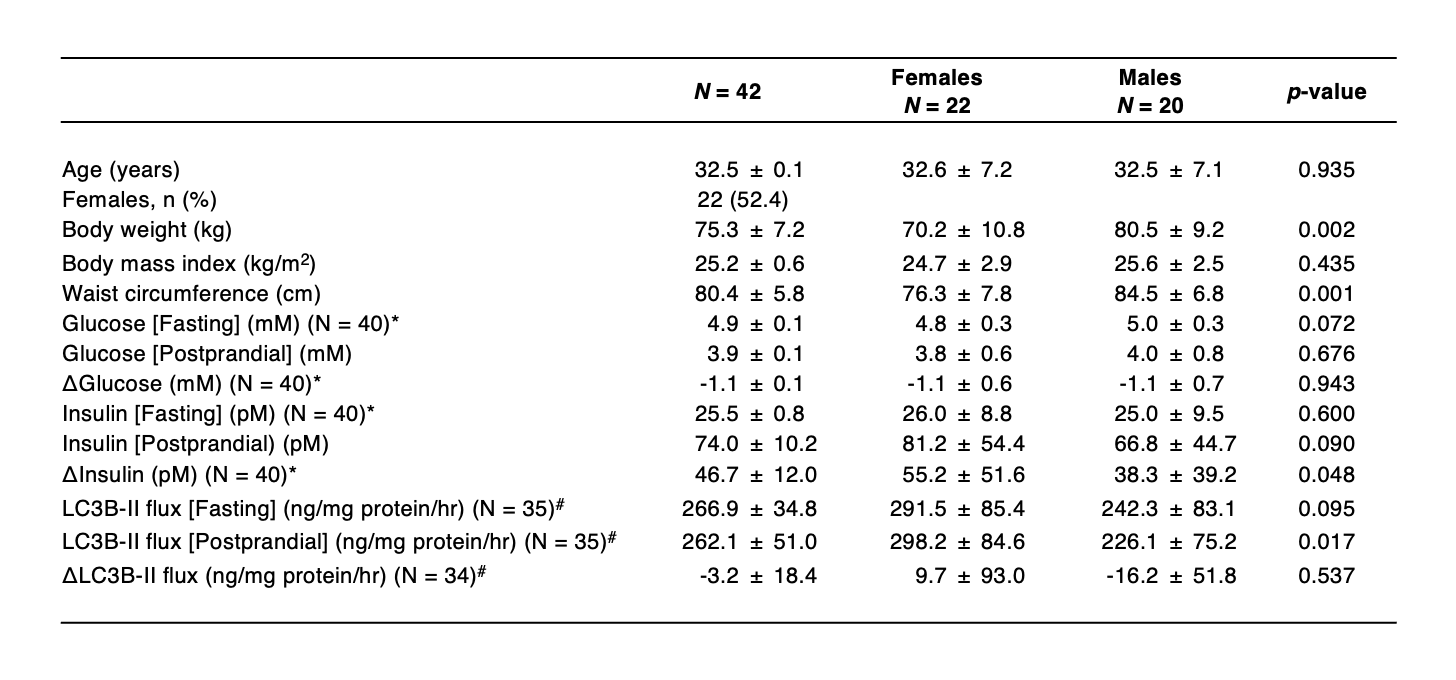
**Table 1 Participant characteristics.** Data are shown as mean ± SD.

^*^ Fasted plasma glucose and insulin measurements missing for 2 participants.

^#^ Autophagy measurements excluded for 6 participants, detailed further under autophagic flux measurement.

**METHODS**

**Study design and setting**

In this report, we present results from 42 participants enrolled in a prespecified, pre-post single arm study (Australian New Zealand Clinical Trials Registry ACTRN12621001029886) between March and September 2022. The study was performed at the South Australian Health and Medical Research Institute (SAHMRI) by researchers from the University of Adelaide and SAHMRI. The inclusion criteria were: aged 20-50 years with a BMI of 18.5-29.9 kg/m^2^. Key exclusion criteria were: co-morbidities likely to affect lysosomal function (e.g. cancer, cardiovascular conditions, neurological disorders) and taking medications capable of altering autophagic activity or metabolism (e.g. blood glucose lowering medications, anti-inflammatories). The detailed study protocol with the complete list of inclusion and exclusion criteria has been published previously [1].

No randomisation was required as each participant served as their own control; all participants provided a pre- and post-intervention blood sample.

**Sex as a biological variable**

Both male and female participants were included in this study, and the effect of sex on autophagy was assessed.

**Intervention**

Participants were instructed to fast overnight for 12 h prior to the study visit. Anthropomorphic data were collected as previously described [1]. A fasting blood sample (20 mL) was drawn. Immediately following blood collection, participants were asked to consume a standardised whey protein drink (30 g unflavoured whey isolate powder diluted in 250 mL skim milk: 842 kJ, Protein 35.25 g, Fats 0.65 g, Carbohydrates 12.2 g) within 5 min. This high-protein, low-carbohydrate drink was expected to increase plasma insulin and reduce plasma glucose, as previously reported [2] – thus inhibiting autophagy via both the insulin and glucose axis. Participants were instructed to remain sedentary for 1 h post-consumption of the high-protein drink, after which a second, postprandial blood sample (30 mL) was collected.

**Outcomes**

The primary outcome of this study was to measure change in autophagic flux after consumption of a protein-rich drink. Secondary outcomes included assessment of changes in plasma glucose and insulin post-consumption of a high-protein drink.

**Autophagic flux measurement**

Autophagic flux was assessed for both fasted and postprandial blood samples immediately following collection, using methodology developed by our group [3,4]. Briefly, 6 mL of blood was divided into 2 tubes – one served as the control, while the other was treated with 150 µM of the lysosomal inhibitor chloroquine (Sigma Aldrich, C6628). Both tubes were incubated for 1 h at 37ºC with rotation. All subsequent steps were performed on ice (or at 4ºC during centrifugation) with cold buffers to inhibit further vesicle trafficking.

Samples underwent a 1:1 dilution in Dulbecco’s Phosphate Buffered Saline (DPBS; Thermo Fisher Scientific, 14190136). PBMCs were isolated by underlaying the blood-DPBS mixture with Lymphoprep (Stemcell Technologies, 07811) and centrifuging for 30 min at 800 x g, with deceleration set to 1. The PBMC layer was collected, diluted in DPBS and pelleted via centrifugation at 600 x g for 10 min. The PBMC pellet was resuspended in 1 mL red blood cell lysis buffer (BD Biosciences, 555899) and incubated on ice for 2 min to remove residual red blood cells. PBMCs were washed twice using DPBS and centrifuged at 600 x g for 10 min. Pellets were resuspended in 1 mL DPBS, transferred to microcentrifuge tubes, and spun at 2,000 x g for 10 min. The supernatant was discarded and PBMC pellets snap-frozen on dry ice and stored at -80ºC.

PBMC pellets were thawed on ice and resuspended in 0.05% saponin (Sigma Aldrich, SAE0073) in DPBS containing protease (Sigma Aldrich, 4693132001) and phosphatase inhibitors (2.5 mM sodium pyrophosphate, 1 mM sodium orthovanadate, 1 mM β-glycerophosphate) for 5 min with gentle rocking. This saponin wash served as a weak permeabilizing agent, removing the cytosolic pool of LC3B-I, without affecting membrane associated LC3B-II, which is what accumulates upon lysosomal inhibition (*i.e.* the species used to quantify autophagic flux). PBMCs were washed in DPBS, re-pelleted and resuspended in cell extraction buffer (Cell Signalling Technology, 35172) containing protease inhibitors. Cell suspensions were sonicated on ice (2x 20 s bursts) and clarified at 16,000 x g for 5 min at 4ºC. The supernatant was collected, and protein concentration determined via a micro-BCA protein assay kit (Thermo Fisher, 23235).

LC3B-II concentration was measured by loading 5 µg clarified cell lysate per well in triplicate on a FastScan Total LC3B ELISA kit (Cell Signalling Technology, 35172). A standard curve was prepared using 0 – 4 ng/mL recombinant human LC3B (Abcam, ab103506). ELISA plates were prepared as per manufacturer’s instructions, and the absorbance measured at 450 nm using Glomax plate reader (Promega). LC3B-II protein concentrations of each sample were interpolated from the standard curve and reported as ng LC3B-II/mg of total protein/hour. Autophagic flux was calculated as:

ΔLC3B-II (ng LC3B-II/mg protein/hour) = LC3B-II [Chloroquine] – LC3B-II [Control]

Autophagic flux was measured in 84 samples (fasting and postprandial samples for n = 42 participants). Two samples (from different participants) were excluded due to coefficient of variation between triplicate wells being > 15%. Twelve samples from the same ELISA plate were excluded due to sample values falling outside the calculated standard curve. Therefore, final autophagic flux analysis was performed on 70 samples from 36 participants.

**Plasma metabolic measurements**

Participant blood was collected in 9 mL EDTA or 2 mL sodium fluoride/potassium oxalate tubes for insulin and glucose measurements, respectively. For glucose measurements, blood was centrifuged at 3000 x g for 15 min at room temperature, with the plasma layer aspirated and stored at -80ºC. Blood glucose was measured using the hexokinase method (Cobas Integra 400 plus, Roche). For plasma insulin measurements, blood was centrifuged at 1500 x g for 10 min at room temperature, after which the plasma layer was aspirated and stored at -80ºC. Plasma insulin was assessed using a commercially available ELISA kit (Mercodia, 10-1113-01).

**Statistics**

Statistical analysis was performed using GraphPad Prism Version 10.1.1 for MacOS. Data distribution was assessed using Shapiro-Wilk test. Missing values were not imputed. Depending on data distribution, either a paired sample t-test or Wilcoxon signed rank test was used to test for differences between fasted and postprandial autophagic flux, plasma glucose and insulin. Spearman’s correlation and two-way ANOVA were used for exploratory analyses. Details for the statistical tests used are provided in the figure legend.

**Study approval**

The study protocol was approved by the University of Adelaide Human Research Ethics Committee (HREC, reference number: H-2021-024). Written informed consent was obtained from all participants.

**Data availability**

Values for all data points used in Figure 1 can be found in the Supporting Data Values file. Other data is available upon reasonable request to the corresponding author.

**ACKNOWLEDGEMENTS**

We acknowledge Xiao Tong Teong for help with glucose measurement. Figure 1A was generated using BioRender.
